# Associations between cardiovascular risk factors, stroke severity, and post-stroke cognition: a moderated mediation analysis

**DOI:** 10.1101/2021.07.05.21260009

**Authors:** Jianian Hua, Yixiu Zhou, Licong Chen, Shanshan Diao, Qi Fang

**Affiliations:** Department of Neurology, The First Affiliated Hospital of Soochow University, Suzhou, Jiangsu, PR China; Medical College of Soochow University, Suzhou 215123, PR China

**Author notes:** Correspondence: Shanshan Diao. Correspondence: Qi Fang. These authors contributed equally (Jianian Hua, Yixiu Zhou).

**Keywords:** stroke, cognitive function, structural equation models, cross-sectional study

## Abstract

**Introduction:** Cognitive impairment may affect one third of the stroke survivors. Cardiovascular risk factors have been described to be risk factors for lower cognition after stroke. However, most previous studies only used multivariate regression models to learn the association. The aim of our study was to investigate whether the effect of cardiovascular risk factors on cognition after stroke was mediated by stroke severity, the estimated effect of direct and indirect pathways, and the moderated association.

**Method:** In this incident cross-sectional study, 300 stroke patients received cognitive test within seven days after stroke. Cognitive tested was performed by the Chinese version of Mini-Mental State Examination (MMSE). A second stage dual moderated mediation model was used the select moderation variables. Finally, we constructed a structural equation model to test the indirect effects of cardiovascular and demographic factors on cognition stroke severity, the direct effects of predictors on cognition, and the moderated effects of hypertension.

**Results:** Age (estimate, -0.114; 95% bias-corrected CI, -0.205, -0.032; P<0.001), female (estimate, -2.196; 95% bias-corrected CI, -4.359, -0.204; P=0.009), lower education (estimate, -0.893; 95% bias-corrected CI, -1.662, --0.160; P<0.001), stroke severity (estimate, -1.531; 95% bias-corrected CI, -3.015, -0.095), hypertension (estimate, -2.242; 95% bias-corrected CI, -4.436, -0.242; P=0.003) and atrial fibrillation (estimate, -4.930; 95% bias-corrected CI, -12.864, -0.126; P=0.048) were directly associated with lower cognitive function after stroke. We found no evidence that cardiovascular risk factors indirectly correlated cognitive function through stroke severity. The combination of hypertension could alleviate the negative effect of atrial fibrillation on cognition (estimate, -3.928; 95% bias-corrected CI, -7.954, 0.029; P=0.009).

**Conclusions:** We explored the complex relationship between cardiovascular risk factors, stroke severity, and cognitive function after stroke. Using our method, researchers using other dataset could repeat the analysis and achieve a better understanding of the relationship. Future researchers are needed to find whether the moderated associations were casual or modifiable.

## 1 Introduction

Nowadays, the incidence rate of ischemic stroke is growing^1^. it is well known that stroke is associated with acute cognitive decline and increased risk of dementia^2,3^. The post-stroke cognitive impairment (PSCI) is an important sequela of stroke. The declined cognition brings great burden to caregivers, economic, and global health^4,5^. Therefore, it is imperative to learn the modifiable risk factors for PSCI.

Recently, cardiovascular disease and stroke severity are of great interest in studies learning PSCI, due to the following reason. They are well-known risk factors for dementia in any of the population^6,7^. Heart and brain communicated intensively regarding to cognition. Patients with stroke had more cardiovascular comorbidity^8^. The previous results are inconclusive or conflicting^9,10^. However, most studies analyzed them through multiple regression, but the interaction relationship and the multiple routes were seldomly learned.

The post-stroke cognitive decline was reported to be composed of incident cognitive decline immediately after stroke and the accelerated pre-stroke cognitive decline^11^. The incident decline might be caused by stroke itself, such as the acute tissue damage, or stroke severity^12^. The pre-stroke cognitive decline was associated with cognitive risk factors which had been existed before stroke. Hence, the mediated model could better explain the above two kinds of cognitive decline. Meanwhile, the moderated model would be used to learn the interaction term. For example, previous studies found the combination of hypertension and diabetes may be detrimental or even beneficial to the cognitive function among stroke patients^13-16^. The possible mechanisms included collateral circulation^17^.

Nowadays, only few studies learned the moderated and mediated effect in the domain of post-stroke cognitive function^12,18-20^. *Drozdowska 2020* studied the effects of cardiovascular disease on PSCI, and they found some risk factors were indirectly associated with the cognition after stroke^19^. However, the association hadn’t been learned in China. Our study was conducted in Soochow, a city located in eastern China, with a life-expectancy over 85 years, which could reflect the medical situation of the population in the developed areas of China. The aim of our study was to explore the complex association between cardiovascular risk factors and the cognitive function in the acute phase after stroke.

## 2 Method

### 2.1 Study design and participants

This study employed a cross-sectional design. We recruited stroke patients who admitted to the stroke unit of the First Affiliated Hospital of Soochow University. Every patient in the stroke unit was provided with standardized treatment, and high dependency clinical and nursing care^21^. During June 2020 and May 2021, three senior vascular neurologists (YX Z, LC C and SS D) collected demographic and clinical data and conducted standardized cognitive assessments for all study patients. The inclusion criteria were: (1) Diagnosis of ischemic stroke was confirmed after admission by MRI^22^; (2) Patients were admitted within seven days of illness. The exclusion criteria were^19,23,24^.: (1) Patients were unable to complete the cognitive test due to existing impairment, such as aphasia. (2) Patients had disturbance of consciousness causing by severe stroke. (3) a history of or current major depression, as determined by clinical reports or PHQ-9 of >9

### 2.2 Standard protocol approvals

All protocols followed those outlined in the Declaration of Helsinki and were approved by Institutional Review Board of The First Affiliated Hospital of Soochow University Hospital (IRB No. 2021-172). In reporting our study, we followed Strengthening the Reporting of Observational Studies in Epidemiology (STROBE) guidelines^25^.

### 2.3 Data availability statement

Data were entered into the stroke registration system of the First Affiliated Hospital of Soochow University (SR-FHSU). Researchers could get data after the approval of the corresponding author and the ethic committee of the hospital.

### 2.4 Predictors

Predictors included demographic factors and cardiovascular risk factors. Demographic factors included age, sex, education, current smoking^6^. Age was treated as a continuous variable. Education level included the following category: primary school or less, middle school, high school and bachelor’s degree or higher. Since almost all patients lived in the city of the hospital, we did not registered living area. Cardiovascular risk factors were those have been associated with post-stroke dementia: hypertension, diabetes mellitus, previous stroke, coronary disease, atrial fibrillation. Due to lack of previous medical record, we did not differentiate previous TIA and previous stroke.

### 2.5 Mediators

Every patient was evaluated by National Institute of Health Stroke Seale (NHISS) on immediately upon admission to our stroke unit.

If the patients arrived in hospital within the time window or tissue window, they would firstly receive intravenous thrombolysis or thrombectomy in the emergency department. Then, they would be admitted to the ischemic stroke unit if they had no hemorrhage. These patients were assessed NHISS score after arriving the stroke unit.

To achieve a more parsimonious model, we categorized NHISS into four groups: no stroke signs (0), minor stroke (1-4), moderate stroke (5-15), and severe stroke (16-42).

### 2.6 Cognitive outcome

Cognitive function was evaluated within their 7-days of ischemic stroke or TIA. It was measured by the Chinese version of Mini-Mental State Examination (MMSE)^26^. We handled both MMSE score as continuous variables.

The MMSE is a widely used tool for the assessment of cognitive function in older participants. It reflects five aspects of cognitive function: orientation, registration, attention and calculation, recall, and language. The total score of MMSE ranges from 0 to 30.

### 2.7 Statistical analysis

As mediation analyses assuming the actual temporal order, we constructed the variables in the following orders. Stroke severity regressed on 11 predictors (including demographic and cardiovascular risk factors). Then, cognitive function regressed on stroke severity, and also regressed on the 11 predictors. It reflected the direct effect of predictors and stroke severity on cognitive function, and the indirect effect of predictors on cognitive function, mediated by stroke severity. To avoid overfitting, we retained all predictors, no matter whether the path was significant^27^.

We developed a second stage dual moderated mediation model. Firstly, we assumed that hypertension and diabetes might moderate the following paths: (1) the direct path between predictor and outcome, and (2) the mediator path between predictor and mediator. That is, namely, to what extent the occurring of hypertension and diabetes (1) change the direct effect of predictors on cognitive function, and (2) change the indirect effect of predictors on cognitive function through stroke severity. We conducted the second stage dual moderated mediation model for atrial fibrillation, coronary disease and previous stroke, separately (Figure 1). For example, while exploring the moderated effect on atrial fibrillation, the model fitted four situations (presence or absence of hypertension × presence or absence of diabetes)^28^. We then explored the moderated effect on other two predictors. Secondly, we removed interaction terms with a P value over 0.20. In the final model (Figure 2), we kept two interaction terms (arrow pointed from hypertension to arrow between atrial fibrillation and cognitive function, arrow between coronary disease and cognitive function).

**Figure 1.**
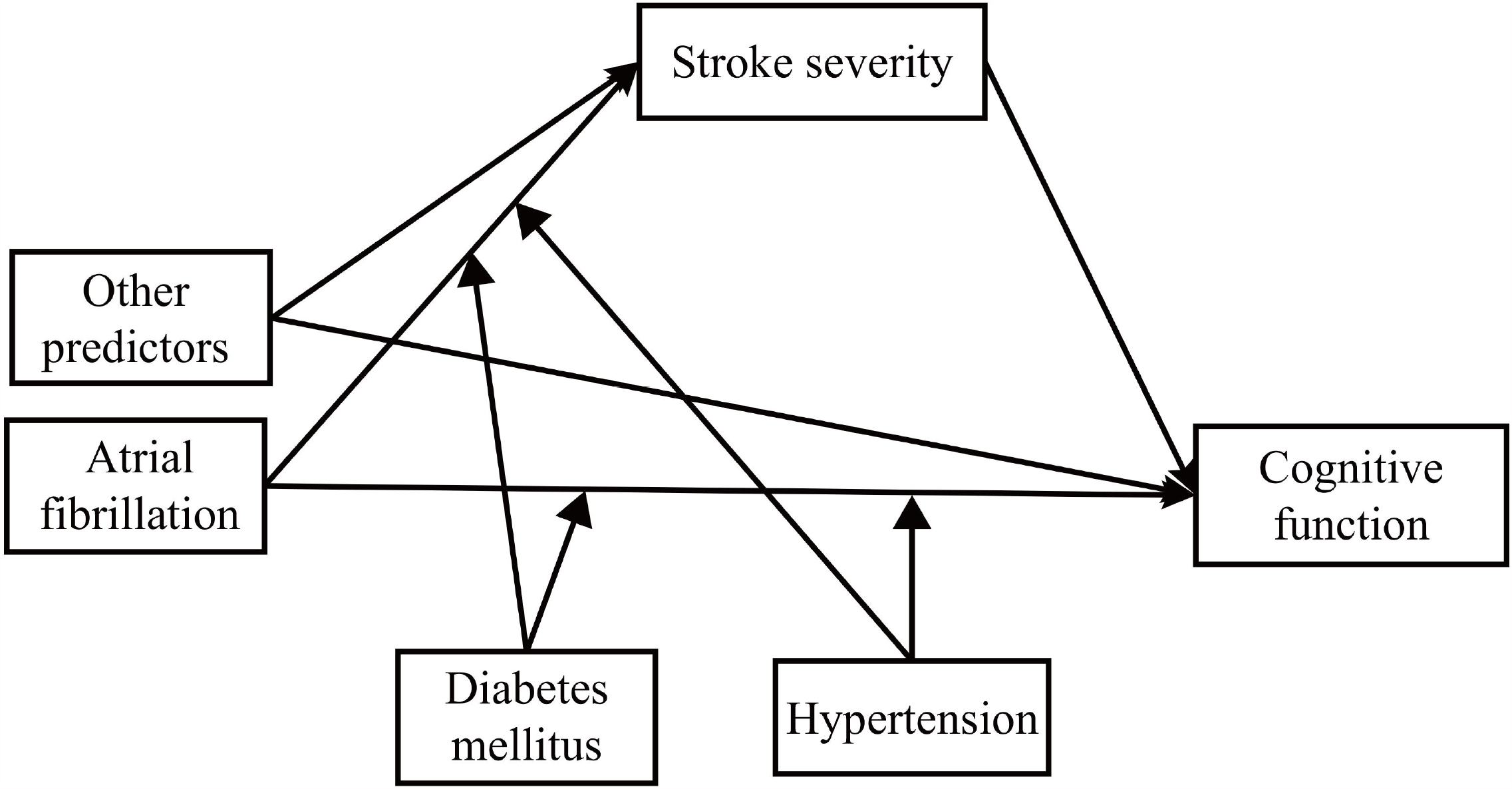
The second stage dual moderated mediation model. We examined the moderated effect on atrial fibrillation, coronary disease and previous stroke, separately.

**Figure 2.**
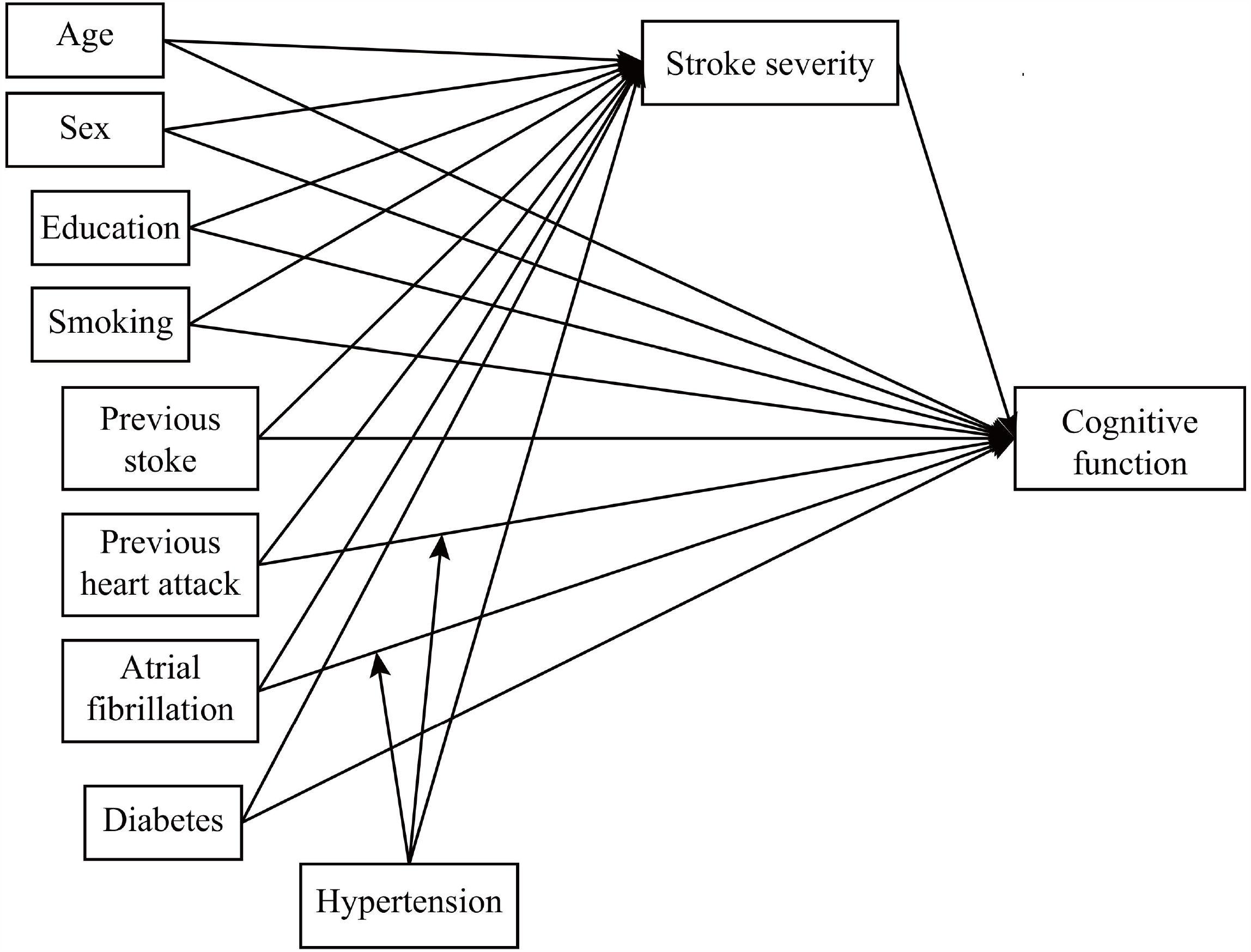
The final diagram of the moderated and mediation model.

The dependent variables were all continuous, therefore, we used the Maximum Likelihood Estimation (ML) method. It relatively would not be constricted to normal distribution and offer better precision for calculating confidence intervals (CI). A 1000-replication bootstrapping was used to estimate the CI^29,30^. The structural equation modelling was performed by Mplus 8.3 (Muthén & Muthén)^31,32^.

## 3 Results

A total of 300 ischemic stroke patients were included in our study. The baseline characteristics were shown in Table 1. The mean±SD age of all participants were 63.5±11.3 years. 74.3% of them were male. 44.6% of the participants hadn’t finished primary school. 21.0% of the participants had a previous history of stroke. The median (interquartile range) of NHISS score was 3 (1, 5). The mean±SD MMSE score of all participants was 17.0±8.6.

**Table 1.**
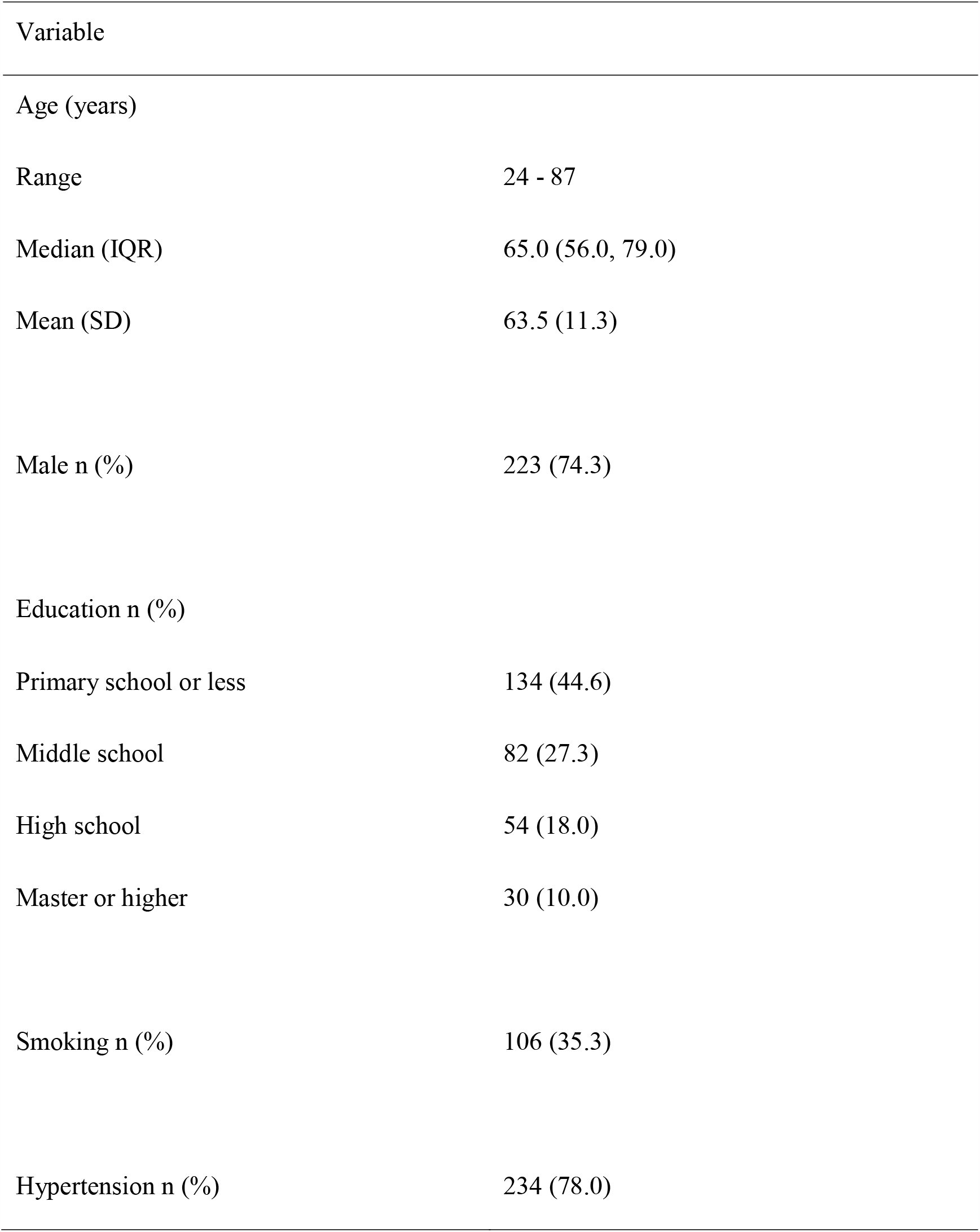
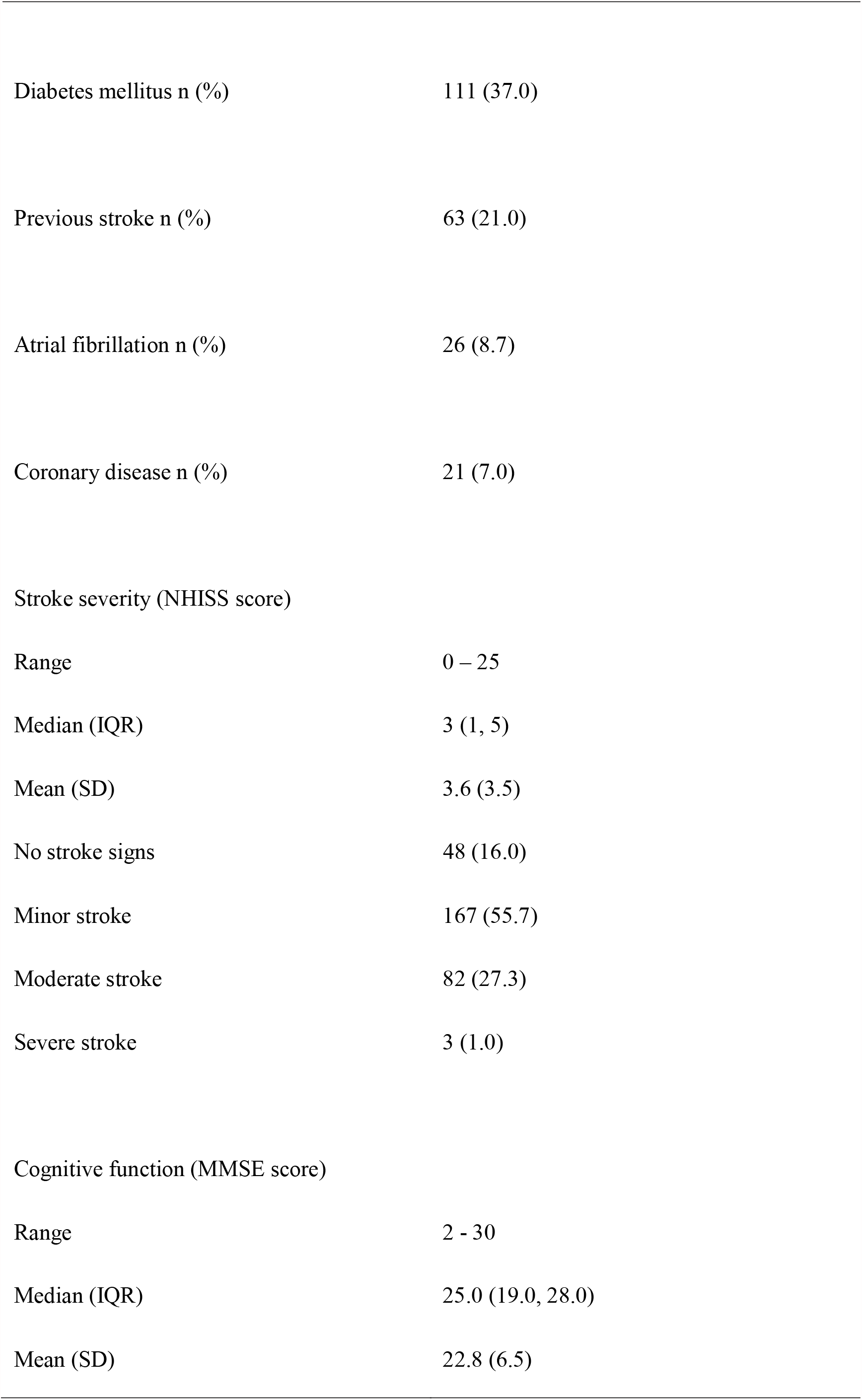
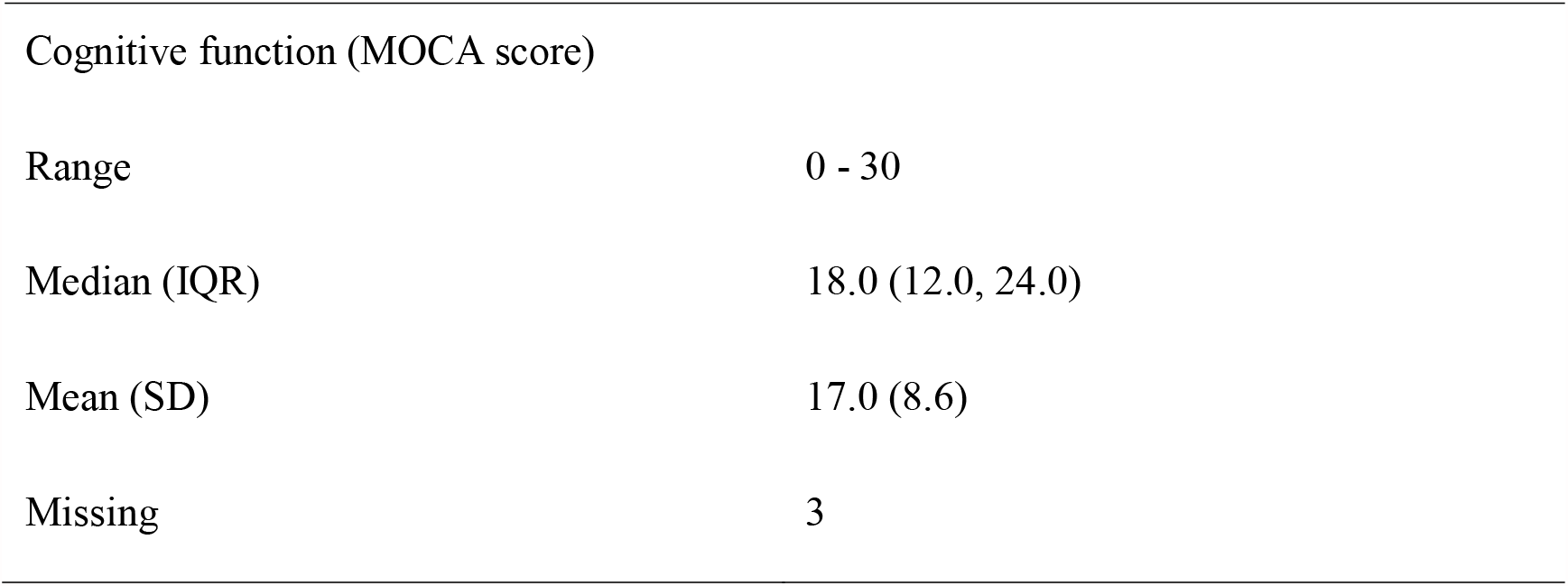
Demographic, clinical and cognitive data of all participants

### 3.1 Final model contracture

Firstly, three individual two-stage moderated mediation models were fitted to evaluated\ the moderation effect. Secondly, we opted to retain two interaction terms in the final model. We considered hypertension as a moderator for effects of atrial fibrillations and coronary disease on cognitive function. The final model was shown in Figure 2. The overall model fitted excellent, with SMSR=0.002.

### 3.2 Mediation effects

As for direct associations, there was a statistically significant association between the mediator and cognitive function. Higher NHISS score was directly associated with lower MMSE score (*β*=-1.531; 95%CI=-3.015, -0.095; P=0.008). Age, sex, education, hypertension and atrial fibrillation were directly related to cognitive function (Table 2). One year of aging was associated with 0.114 points decrease in MMSE score (*β*=-0.114; 95%CI=-0.205, -0.032; P<0.001). Male had high MMSE score (*β* =-2.196; 95%CI=0.204, 4.358; P=0.009). High education was associated with high score (*β*=0.893; 95%CI=0.160, 1.662; P<0.001). Patients with hypertension or had lower score (*β*=-2.242; 95%CI=-4.436, -0.242; P=0.003). Atrial fibrillation was also correlated with lower score (*β*=-4.930; 95%CI=-12.864, -0.126; P=0.048). There existed a trend that participants with coronary disease had higher cognitive function (*β*=2.193; 95%CI=-1.103, 4.303; P=0.048).

**Table 2.** Direct associations between predictors and cognitive function

|  | Unstandardized coefficient (95% CI) | P value |
| --- | --- | --- |
| Age | -0.114 (-0.205, -0.032) | <0.001 |
| Sex (male) | 2.196 (0.204, 4.359) | 0.009 |
| Education | 0.893 (0.160, 1.662) | <0.001 |
| Smoking | 0.664 (-1.198, 2.629) | 0.378 |
| Hypertension | -2.242 (-4.436, -0.242) | 0.003 |
| Diabetes mellitus | -0.120 (-2.013, 1.651) | 0.870 |
| Previous stroke | -0.992 (-3.344, 1.238) | 0.259 |
| Atrial fibrillation | -4.930 (-12.864, -0.126) | 0.048 |
| Coronary disease | 2.193 (-1.103, 4.303) | 0.054 |
| Atrial fibrillation<br>× hypertension | -3.928 (-7.954, 0.029) | 0.009 |
| Coronary disease<br>× hypertension | 3.195 (-1.719, 8.482) | 0.106 |

For indirect associations, we observed no significant indirect effects of predictors on cognitive function (Table 3). None of the predictors were correlated with more severe strokes (Supplementary Table 1).

**Table 3.** Indirect associations between predictors and cognitive function

|  | Unstandardized coefficient (95% CI) | P value |
| --- | --- | --- |
| Variable |  |  |
| Age | 0.001 (-0.002, 0.040) | 0.149 |
| Sex (male) | 0.006 (-0.456, 0.463) | 0.967 |
| Education | 0.079 (-0.027, 0.297) | 0.185 |
| Smoking | -0.144 (-0.744, 0.218) | 0.381 |
| Hypertension | 0.285 (-0.049, 1.240) | 0.158 |
| Diabetes mellitus | -0.085 (-0.596, 0.238) | 0.550 |
| Previous stroke | 0.016 (-0.448, 0.634) | 0.928 |
| Atrial fibrillation | -0.313 (-1.237, 0.252) | 0.242 |
| Coronary disease | -0.183 (-1.127, 0.335) | 0.438 |

### 3.3 Moderation effects

The direct effects of atrial fibrillation and coronary disease could be moderated by hypertension. Co-occurring with hypertension, atrial fibrillation was associated with higher cognitive function (*β* changed from -4.930 to -3.928). Through probing, we observed hypertension had a positive effect on cognitive function among patients with coronary disease (*β*=3.195; 95%CI=-1.719, 8.482; P=0.106).

## 4 Discussion

Using a real-word sample of stroke unit patients, this article learned the relationship between cardiovascular risk factors, stroke severity and cognition. We found age, sex, education, atrial fibrillation, hypertension, and stroke severity were directly associated with lower cognitive function during the acute phase after stroke. For patients with atrial fibrillation or coronary disease, there was a trend that a combination hypertension was associated with better cognitive function. We also found a trend that patient with coronary disease probably had higher cognitive function.

Our findings were partly different from the study of *Drozdowska 2020*^*19*^. Drozdowska thought age and atrial fibrillation indirectly correlated with lower cognitive function through stroke severity. We found age, female, lower education, atrial fibrillation and hypertension was directly associated with cognitive function. In our dataset, the predictors were not associated with the mediator (stroke severity). The significance of indirect association needs both the significance of “predictor to mediator” and “mediator to outcome”. Moreover, Drozdowska hadn’t learned the effect of education. In Drozdowska’s country, England, sex was not associated with cognition. In China, older female had lower cognitive function, which had already supported by our previous nation-wide cohort study and many other articles^33,34^. In agreement with Drozdowska, we found a trend that hypertension had a positive effect on cognition among participants with coronary disease. We also found the positive effect of hypertension on patients with atrial fibrillation.

Several mechanisms could explain our findings. Hypertension was associated with lower cognitive function. It could bring with remodeling of cerebral vessels, lipohyalinosis, atherosclerosis, and oxidative stress. The pathological mechanism would in turn lead to cognitive decline, and other neurological dysfunctions^35^. Atrial fibrillation was independently associated with lower cognitive function, among participants with or without stroke^36-38^. A review discussed AF may result in a serious of mechanism that would cause lower cognition, such as cerebral hypoperfusion, inflammatory responses, silent ischemia, reduced brain volumes, and cerebral microbleeds. A combination of hypertension might reverse the effect of cerebral hypoperfusion, and therefore alleviated the negative effect. This was the potential explanation of our finding in the moderated analysis. It is widely reported that AF was associated with higher severity of stroke^39,40^. However, in our dataset, AF was not associated with NHISS score. We assumed this might be due to “survival bias”. Sever stroke patients with AF was unable to finish the cognitive test, and thus was excluded from the research. This also explains why also explain why the incidence rate of AF was lower than other stroke datasets (about 17%). Notably, it was reasonable that other predictors were not correlated with stroke severity^19,40^. The association between coronary disease and cognitive function is still controversial^41,42^. The combination of higher blood pressure would bring with better collateral flow and better clinical outcomes^43^. This could explain the positive effect in our article. Although during the acute phase of stroke, high blood pressure would cause increasing infarct volume, brain edema, and hemorrhagic transformation. Every patient in our stroke unit received standardized treatment, including blood pressure control^44^.

To our knowledge, this was the first study learning the moderated and mediated association between cardiovascular risk factors, stroke severity, and cognitive function in among Chinese stroke patients. We believed that extensive researchers of the interaction between risk factors would achieve a better understanding of the neuro-degenerative process. However, we also had several disadvantages. Firstly, our data set had more males than females. This was due to the patient distribution of the hospital and district^45^. In the future research, we would try our best to improve this distribution. Second, our dataset included a few sever stroke patients. However, sever stroke patients always had problem in consciousness and language function. The disability prevented us from doing cognitive test. It has always been an intractable problem for studies learning cognitive function or dementia. Third, we could not distinguish previous TIA from previous stroke. We were not able to get the previous medical record of our patients, which were not admitted to our hospital before. Furthermore, in our district, patients had relative lower education level, and thus, had lower awareness rate of the previous history.

## 5 Conclusions

Our research explored the complex relationships between cardiovascular risk factors, stroke severity, and cognitive function. We highlighted the importance of comorbidity while learning the cognitive function after stroke. Future studies are needed to learn whether our findings are casual.

## 7 Author Contributions

Qi Fang and Jianian Hua contributed to the conception and design of the study. Shanshan Diao, Yixiu Zhou and Licong Chen performed cognitive tests and collected medical data. Jianian Hua performed the statistical analysis. Jianian Hua wrote the first draft of the manuscript. Qi Fang reviewed the manuscript. All authors approved the final version of the paper.

