## Supplementary Materials for "Associations between cardiovascular risk factors, stroke severity, and post-stroke cognition: a moderated mediation analysis"

Supplementary Material

Table S1 Direct associations between predictors and mediators.

|  | Unstandardized coefficient (95% CI) | P valure |
| --- | --- | --- |
| Age | -0.007 (-0.020, 0.002) | 0.068 |
| Sex (male) | -0.004 (-0.232, 0.238) | 0.964 |
| Education | -0.051 (-0.130, 0.021) | 0.084 |
| Smoking | 0.093 (-0.147, 0.340) | 0.314 |
| Hypertension | -0.185 (-0.430, 0.058) | 0.060 |
| Diabetes mellitus | 0.005 (-0.173, 0.257) | 0.497 |
| Previous stroke | -0.010 (-0.297, 0.245) | 0.921 |
| Atrial fibrillation | 0.203 (-0.235, 0.621) | 0.188 |
| Coronary disease | 0.118 (-0.288, 0.459) | 0.393 |
